# Ethnic variation and structure-function analysis of tauopathy-associated *PERK* alleles

**DOI:** 10.1101/2024.03.03.24303689

**Authors:** Goonho Park, Angela Galdamez, Keon-Hyoung Song, Masako Le, Kyle Kim, Jonathan H. Lin

**Author notes:** To whom correspondence should be addressed: 300 Pasteur Dr. L235, Stanford, CA 94305.

## Abstract

EIF2AK3, also known as PERK, plays a pivotal role in cellular proteostasis, orchestrating the Unfolded Protein Response (UPR) and Integrated Stress Response (ISR) pathways. In addition to its central position in intracellular stress regulation, human GWAS identify EIF2AK3 as a risk factor in tauopathies, neurodegenerative diseases caused by aberrant tau protein accumulation. Guided by these genomic indicators, our investigation systematically analyzed human PERK variants, focusing on those with potential tauopathy linkages. We assembled a comprehensive data set of human PERK variants associated with Wolcott Rallison Syndrome (WRS), tauopathies, and bioinformatically predicted loss-of-function, referencing the gnomAD, Ensembl, and NCBI databases. We found extensive racial/ethnic variation in the prevalence of common *PERK* polymorphisms linked to tauopathies. Using SWISS-MODEL, we identified structural perturbations in the ER stress-sensing luminal domain dimers/oligomers of tauopathy-associated PERK variants, Haplotypes A and B, in combination with another tauopathy-linked R240H mutation. Recombinant expression of disease-associated variants *in vitro* revealed altered PERK signal transduction kinetics in response to ER stress compared to the predominant non-disease variant. In summary, our data further substantiates that human PERK variants identified in tauopathy genetic studies negatively impact PERK structure, function, and downstream signaling with significant variations in prevalence among different racial and ethnic groups.

## INTRODUCTION

Eukaryotic translation initiation factor 2 alpha kinase 3 (EIF2AK3), commonly known as Protein kinase R-like endoplasmic reticulum kinase (PERK), is an important regulator of the unfolded protein response (UPR) and integrated stress response (ISR). PERK regulates translation to maintain cellular proteostasis during periods of ER stress[1–3]. PERK senses ER stress through its luminal domain, while the kinase domain phosphorylates the eIF2α subunit to attenuate mRNA translation during stress conditions[1,4].

In humans, genetic studies identified PERK as a causal disease gene or risk factor in Wolcott-Rallison syndrome (WRS)[5]; Progressive Supranuclear Palsy (PSP)[6–8]; and some forms of Alzheimer’s Disease (AD)[9,10]. These diseases are associated with distinct genetic variants in the human PERK gene that impact its function. For example, cells expressing the tauopathy-associated Haplotype B PERK variant show reduced kinase activity coupled with increased vulnerability to ER stress-induced damage[11,12].

Our previous research examined the functional consequences of tauopathy-associated PERK variants categorized into protective Haplotype A and risk Haplotype B, which showed significantly diminished kinase activity in response to ER stress. We investigated the underlying mechanisms of this reduced activity, finding that the Haplotype B variants (S136C, R166Q, and S704A) including D566V variants within the cytosolic kinase domain led to decrease in the protein downstream signaling activity, and the S136C polymorphism specifically caused abnormal disulfide bridging and increased protein turnover. Additionally, we identified that PSP patient iPSC-derived neurons carrying tauopathy-associated hypomorphic PERK alleles had decreased PERK activity in response to ER stress, heightened sensitivity to apoptosis, and increase in total and phosphorylated tau levels. Pharmacological inhibition of PERK also led to increased neuronal cell death and tau protein dysregulation, reinforcing the vital role of PERK signaling in neuronal survival and protein homeostasis under stress conditions[11].

More recently, we found that tauopathy-related variants, especially Haplotype B, showed a higher prevalence in East Asian populations, suggesting a potential race-specific impact on PERK’s function. In addition, we found that the R240H variant identified from genetic studies in Dutch Alzheimer’s disease patients negatively affected the PERK activity in bioinformatic modeling analysis. Structural modeling of tauopathy-linked PERK variants indicated that Haplotype B and the R240H polymorphisms disrupted crucial hydrogen bonds in the luminal domain, potentially altering PERK’s function and stability. Our data supports that many tauopathy-linked PERK variants carry an altered stress-sensing luminal domain but preserved kinase function[12].

Here, we further investigate structural and functional impacts of tauopathy-associated PERK mutations using computational modeling and experimental comparison of signaling activities of recombinant PERK variants expressed in *PERK^-/-^* mouse embryo fibroblast (MEF) cells.

## RESULTS

### Genomic profiling of disease-associated human PERK mutations

In our study, we initially focused on elucidating predicted pathological effects of human PERK mutations. The comprehensive diagram of the PERK protein, as presented in Figure 1, distinctly illustrates amino acid alterations. These alterations are associated with Wolcott-Rallison syndrome (Fig. 1A), tauopathies (Fig. 1B), and predicted loss-of-function (pLoF) variants (Fig. 1C), as sourced from gnomAD, NCBI, Ensembl, and PubMed literature[13].

**Figure 1:**
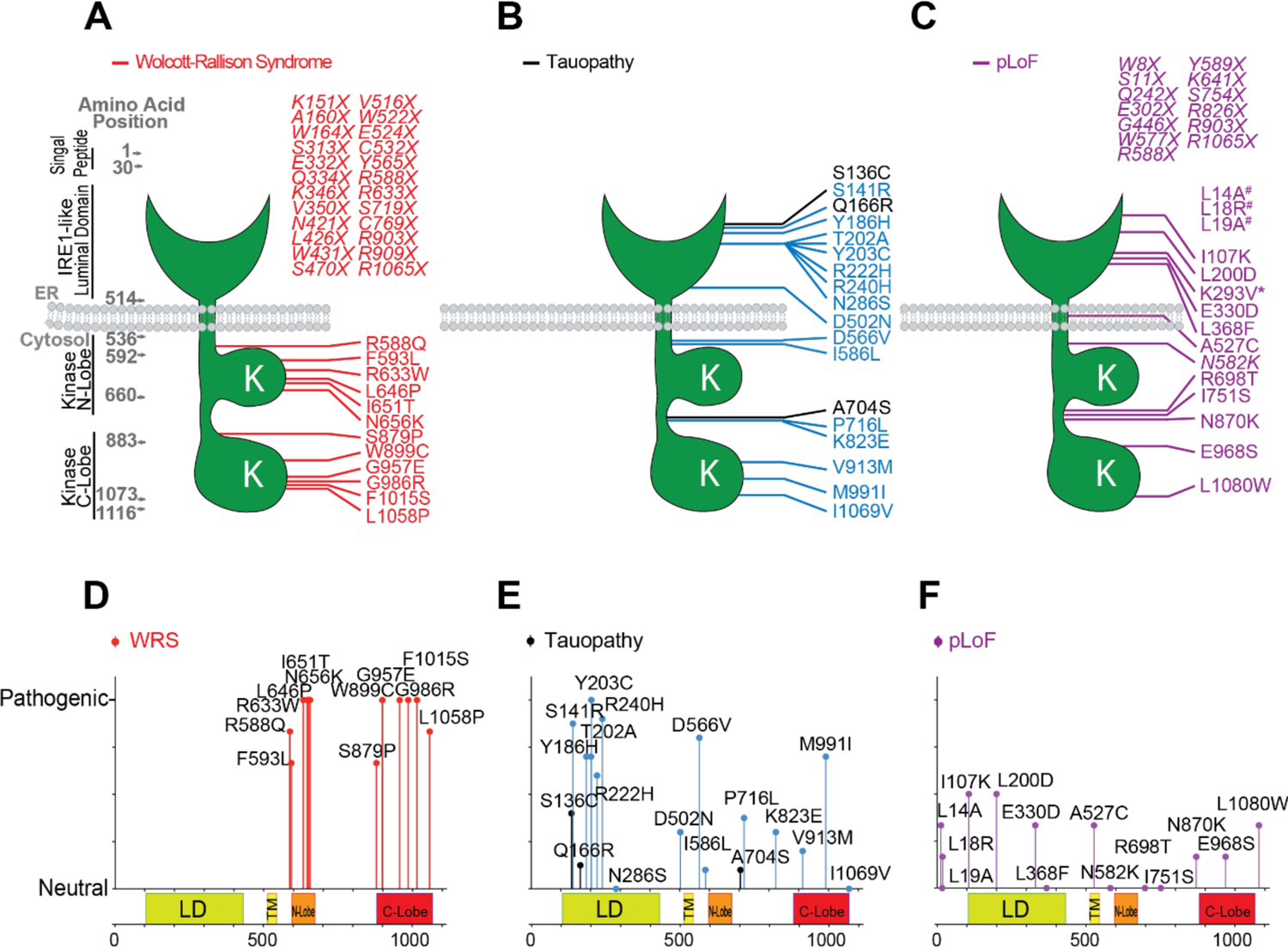
Summary illustration of pathological human PERK mutations. The diagram of the PERK protein highlights the amino acid alterations associated with (A) Wolcott-Rallison syndrome (WRS), (B) tauopathies, and (C) predicted loss of function (pLoF), as sourced from gnomAD, NCBI databases, and prior literature. Color-coded lines (red, blue, black, and purple) represent missense mutations. The symbol (X) designates a stop codon that results in a prematurely terminated protein form. Mutations depicted in black are those with a common occurrence, characterized by an allele frequency greater than 1%. In contrast, mutations shown in blue are considered rare, with an allele frequency less than 1%. WRS and pLoF alleles are categorized as having a rare frequency. Figures 1D, 1E, and 1F depict a lollipop graph detailing the pathogenic significance of individual missense mutations, derived from analyses using five distinct bioinformatics tools. Key functional domains of the 1116-amino acid PERK protein - including the IRE1-like Luminal Domain (LD), the ER transmembrane domain (TM), as well as the N-terminal (N-lobe) and C-terminal (C-lobe) kinase regions in the cytoplasm - are shown.

Interestingly, most missense mutations associated with Wolcott-Rallison syndrome (WRS) are located within the two cytosolic kinase domains[12,13] and exhibit a high degree of pathogenicity using combinational analysis of 5 bioinformatic tools: PolyPhen-2, PROVEAN, MutationTaster, SIFT, and CADD (Fig. 1D). On top of that, 24 WRS-related nonsense mutations are dispersed throughout the PERK gene likely leading to no stable protein production. In the context of tauopathies, no nonsense mutations have been reported. However, missense mutations linked with tauopathies that display higher pathogenicity are predominantly situated in the luminal domain. Furthermore, in our assessment of 43 unique PERK pLoF variants listed in the gnomAD database, we excluded 7 variants deemed low confidence by the gnomAD quality control process and another 8 variants categorized as silent or low-quality pLoF mutations. This refinement led to the identification of 28 high-confidence pLoF variants distributed throughout PERK, including 13 nonsense variants and 15 missense changes. Interestingly, the pathogenic impact of these 15 missense pLoF mutations was relatively mild compared to that of WRS and tauopathy missense variants (Fig. 1D, 1E, and 1F). Ultimately, our analysis supports that WRS variants cause greater damage than tauopathy variants and identifies additional pLoF changes as candidate disease variants.

### Ethnic distribution and allele frequency of PERK SNPs

In this study, we further explored the racial and ethnic disparities in allele frequencies of PERK SNPs linked to Haplotype B, which are implicated in the risk of tauopathies. We analyzed individual SNP variants—namely S136C, Q166R, A704S, and c-8A>G, collectively known as PERK Haplotype B variants. Our examination utilized data from gnomAD and spanned 8 distinct racial and ethnic groups. The analysis highlighted substantial variations, as detailed in Figures 2A and 2B. The allele frequencies of Q166R, A704S, and c-8A>G showed similarity across different races, contrasting with the variant S136C, which exhibited an opposite frequency pattern. Figure 2C further provides an intricate depiction of the distribution prevalence of these variants across different racial and ethnic groups. The R-correlation graph in Figure 2D reveals an interesting correlation pattern: a positive correlation (indicated in blue) is observed among c-8A>G, Q166R, and A704S, while a negative correlation (shown in red) is evident with S136C in human PERK variations. In conclusion, this study confirmed that racial and ethnic differences in PERK gene variants linked to tauopathies[12], finding notable variations in allele frequencies and correlations among different groups.

**Figure 2:**
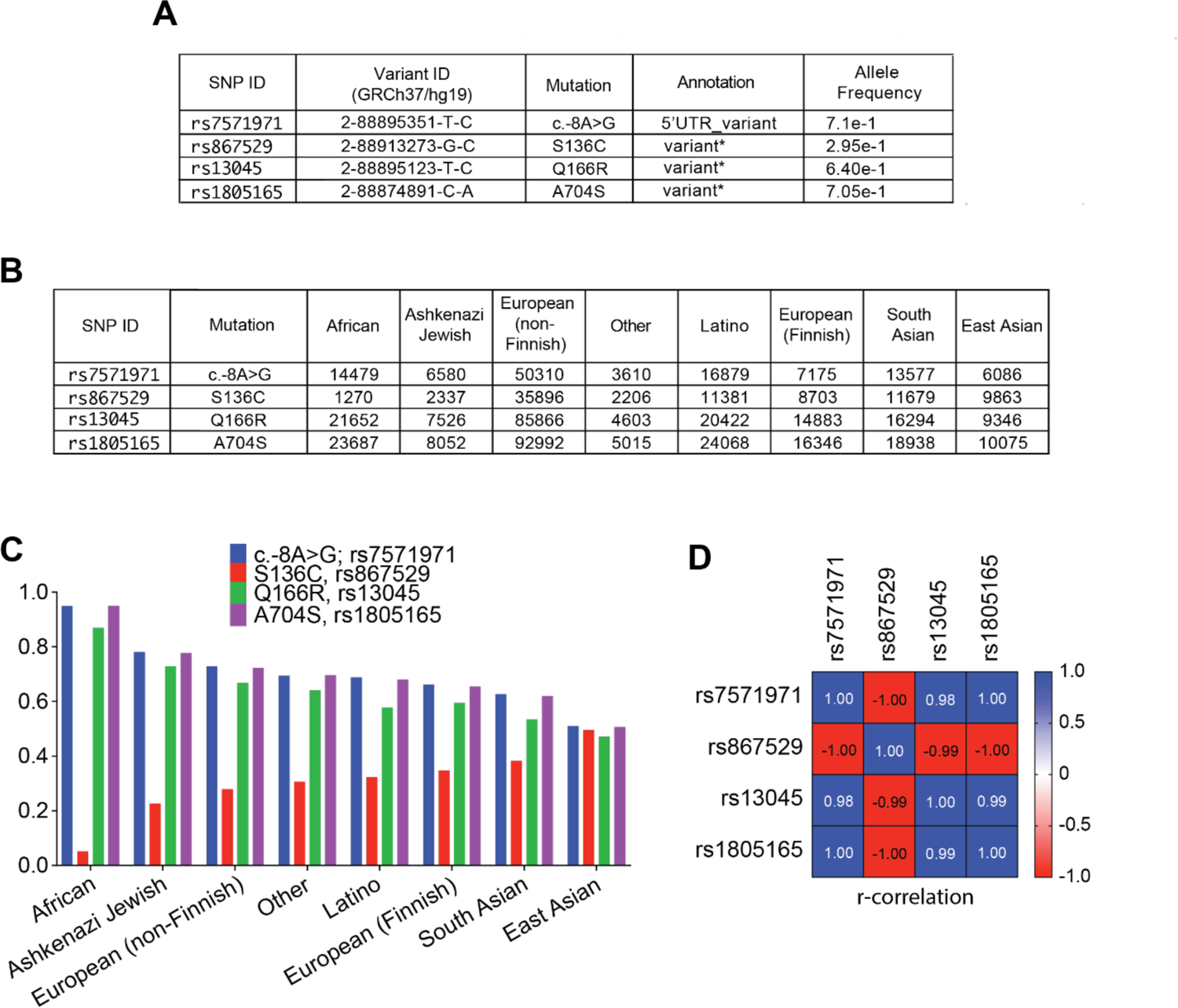
Ethnic distribution and allele frequency for Haplotype B-related PERK SNPs. (A & B) Comparison of total allele frequencies and ethnic populations for PERK SNPs associated with Haplotype B among 8 unique racial/ethnic groups, based on gnomAD data. (C) Graphic representation of the prevalence of these PERK variations across the 8 distinct racial/ethnic segments. (D) R-correlation showcases positive correlations in blue and negative ones in red for the quartet of human PERK variations.

### Three-dimensional (3D) structural insights into the PERK luminal domain

Through structural modeling, previously, we found that tauopathy-associated Haplotype B and R240H PERK variants lost hydrogen bonds in the ER stress-sensing PERK luminal domain[12]. Here, we used SWISS-MODEL to explore how protective and risk ER luminal domain PERK variants affected dimerization and tetramerization structures. We modeled the sequences of PERK Haplotype A as the protective allele in contrast to Haplotype B as the risk allele in combination with the R240H mutation identified in Dutch Alzheimer’s disease patients that also impacts PERK’s luminal domain[10].

Notably, when comparing the dimerization of the protective R240 variant combined with PERK Haplotype A to the tauopathy-associated H240 variant, we found that the H240 mutation induced alterations in the side loop domains (compare L1, L2, and L4 in Fig. 3A) but did not change the beta-sheets and alpha-helices crucial for the internal dimerization interface. Interestingly, no structural differences in loop domains or dimerization interfaces were seen when the R240H alteration was modeled with the PERK Haplotype B variant (Fig. 3B).

**Figure 3:**
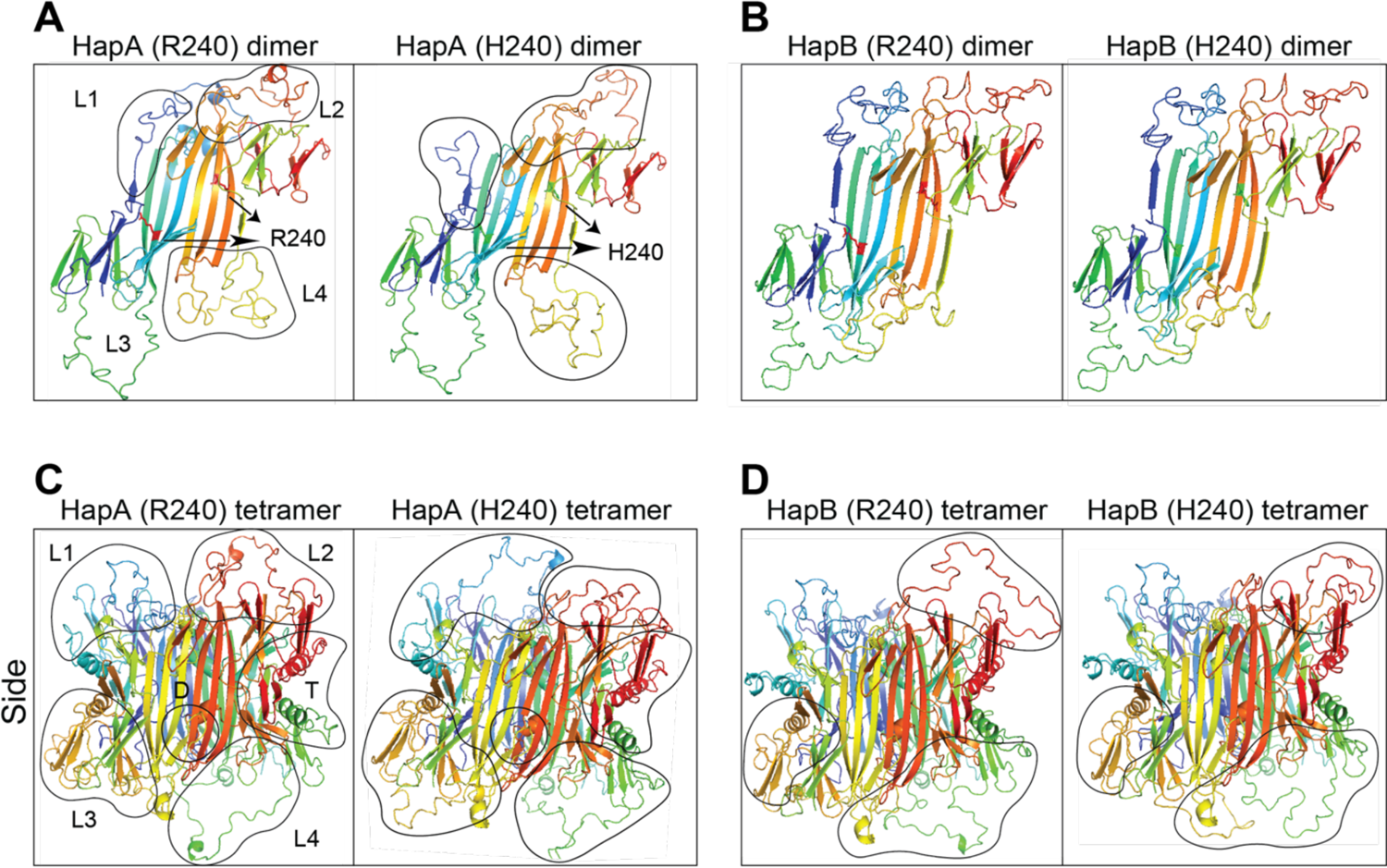
Modeling ER luminal domain of pathogenic PERK variants on three-dimensional protein structure using SWISS-MODEL. (A) The protective R240 variant of PERK Haplotype A in its dimeric form is contrasted with the tauopathy-linked H240 variant, with significant alterations (black circles). (B) Similarly, the protective R240 variant of PERK Haplotype B is compared to its H240 risk variant in dimer form. The tetrameric structures of R240 from Haplotype A (C) and H240 from Haplotype B (D) are showcased, with changes in the luminal domain highlighted by black circles. Arrowheads indicate R240 (red) and H240 (green). D: dimerization interface; T: tetramerization subdomain.

Next, we modeled the impact of these variants upon the tetrameric structure of the PERK luminal domain (Fig. 3C and 3D). A standout observation was the extensive changes caused by the R240H mutation across various loop domains, including the dimerization interface (D) and the tetramerization domain (T) in PERK Haplotype A (Fig. 3C). However, these tetrameric alterations were notably reduced in the PERK Haplotype B variant (Fig. 3D). In summary, our modeling studies identify structural changes in dimerization and tetramerization caused by pathological PERK luminal domain variants.

### ER Stress-induced signaling sensitivity in PERK Haplotypes A and B

Our bioinformatic and structural modeling studies suggested that PERK Haplotype B and the R240H variant altered PERK function. Indeed, we previously demonstrated that PERK Haplotype B had reduced eIF2α phosphorylation activity compared to Haplotype A when expressed in *PERK^-/-^*MEFs and in patient iPSC-derived neurons carrying these variants[11].

Here, we cloned four expression constructs of PERK Haplotypes A and B in combination with the R240H mutation and expressed them in *PERK^-/-^* MEFs. We found reduced phosphorylation of eIF2α by Haplotype B carrying R240 compared to Haplotype A in response to increasing amounts of thapsigargin-induced ER stress (Fig. 4A and 4B), consistent with our prior studies[11]. We monitored PERK protein activation through its migration shift on gels as a response to ER stress, which is indicative of PERK phosphorylation upon activation (Fig. 4A, 4C, 4E, 4G, and 4I). Furthermore, the R240H conversion in Haplotype A led to a decrease in eIF2α phosphorylation in response to ER stress (Fig. 4C and 4D). By contrast, we saw no significant differences in eIF2α phosphorylation by Haplotype B when arginine 240 was converted to histidine in ER stress dose-response experiments (Fig. 4E and 4F). These findings support that the R240H PERK variant encodes a functional hypomorph in the context of the predominant Haplotype A allele. Interestingly, when we challenged cells expressing the R240H variant to high concentrations of thapsigargin, we saw increased eIF2α phosphorylation by the hypomorphic variant (Fig. 4G, 4H, 4I, and 4J), suggesting dysregulation of PERK signaling at both low and high ER stress levels. Alternatively, increased eIF2α phosphorylation at high doses of thapsigargin could reflect the activity of other eIF2α kinases, such as GCN2, which has been reported to phosphorylate eIF2α under conditions of high of ER stress[14].

**Figure 4:**
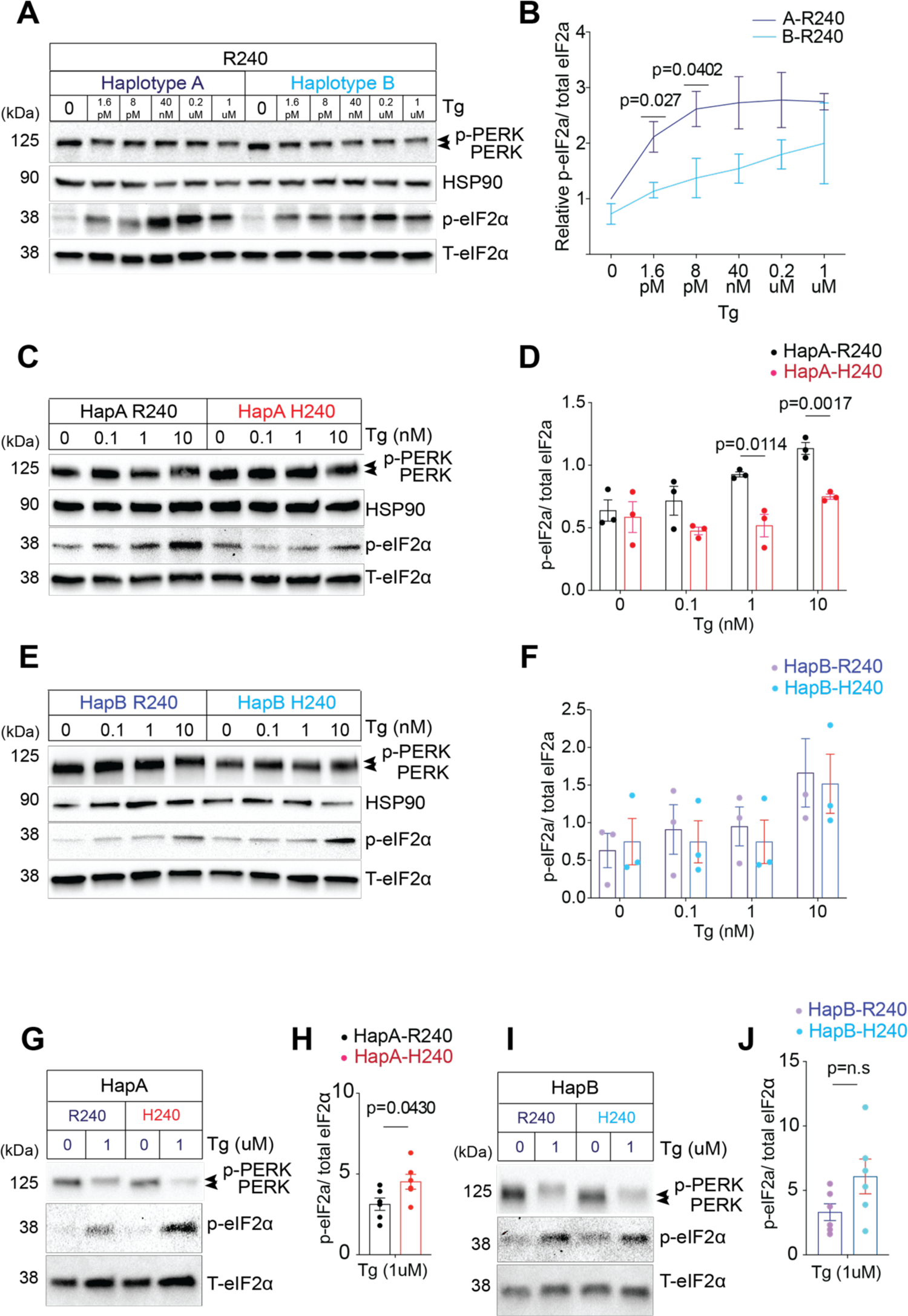
Western blot analysis of ER stress-induced signaling sensitivity in PERK Haplotypes A & B with the R240H mutation (A, C, E, G, and I). Immunoblot and statistical assessment of the protective R240 variant distinguishing Haplotypes A and B linked to human tauopathy using markers, HSP90, PERK, and total (T)-eIF2α for loading control and phosphorylated (p)-eIF2α for ER stress sensitivity. The migration shift of phosphorylated (p)-PERK is observed in total PERK’s western blot. (C & D) Immunoblot and statistical assessment of Haplotype A’s protective R240 and the risk-associated H240 variants, illustrating changes in downstream signaling due to a serial low concentration of thapsigargin. (E to J) Immunoblot and statistical assessment of Haplotypes A and B with the R240H mutation under a serial high concentration of thapsigargin. Error bars show SEM for n=3. The significance of the p-value results is determined by two-tailed Student’s t-test.

## DISCUSSION

Previously, we identified structural and functional consequences of tauopathy-associated PERK variants, Haplotype B and the D566V conversion, on ER stress signaling. Here, we extended these investigations to identify additional potential pLoF PERK disease variants. We confirmed previous racial and ethnic disparities in PERK Haplotype B frequency at 4 individual SNPs[12]. Through three-dimensional structural modeling, we reveal how another tauopathy PERK variant, R240H, can induce structural shifts in PERK dimerization and tetramerization. Last, we provide experimental support that the bioinformatically and structurally predicted changes caused by R240H alter PERK signaling kinetics. These findings lend support that tauopathy EIF2AK3 risk variants functionally impair PERK signal transduction. Future studies will elucidate how reduced or altered PERK activity leads to tau aggregation and neurodegeneration.

Our investigation into the pathogenic variations of PERK shed light on critical aspects of Wolcott-Rallison syndrome (WRS) and tauopathies, incorporating loss-of-function (pLoF) variants which are instrumental in genetic evaluation. A finding from our study is the locational specificity of these mutations within the PERK protein; missense mutations are predominantly situated within the kinase domains in WRS, consistent with increased pathogenicity and negative impact on enzymatic activity (Fig. 1A). Conversely, tauopathies are characterized by a higher frequency of missense mutations within the luminal domain, suggesting their change in ER stress detection and thereby activation kinetics (Fig. 1B). While pLoF missense variants found throughout PERK showed mild pathogenic effects, the proximity of certain mutations, such as L200D in pLoF, T202A & Y203C in tauopathies, N870K in pLoF, and S879P in WRS, suggest these regions are critical for PERK’s activity.

Our genomic study revealed an intriguing population distribution of PERK Haplotype B variants, marked by certain single nucleotide polymorphisms (SNPs), which are predominantly observed in East Asian groups, particularly Japanese (Fig. 2B). This aligns with the increased Progressive Supranuclear Palsy (PSP) incidence in Japanese populations[15,16]. Multiple GWAS identified PERK as a risk factor in the development of PSP in prior literature[6–8]. However, a 2018 GWAS meta-analysis study did not replicate EIF2AK3 as a PSP risk factor[17]. We suggest this inconsistency might be explained by the natural variation in the prevalence of PERK Haplotype B among different ethnicities, since in the 2018 GWAS meta-analysis, multiple diverse racial groups sequencing data for both control and disease cohorts were combined for the meta-analysis. Further research is needed to elucidate why PERK Haplotype B distribution varies across race/ethnicity.

In prior work using structural modeling, we found that tauopathy-related mutations at residues 136 and 240 in the PERK luminal domain disrupt hydrogen bonding within individual PERK monomeric protein. This disruption may be critical for maintaining the monomeric PERK protein’s tertiary structure. While protective variants of PERK could form seven potential hydrogen bonds, disease-associated mutations at these residues compromise their formation, potentially affecting the protein’s ER stress-sensing abilities. Five bioinformatic prediction tools (PolyPhen-2, PROVEAN, MutationTaster, SIFT, and CADD) also suggested that these mutations are pathogenic, with both R240H and S136C identified as risky variants in these algorithms. In our current study, we further identified alterations in the dimer or tetramer PERK structures with the introduction of PERK disease variants. A further computational structural modeling of PERK Haplotype B and the Dutch Alzheimer’s Disease mutation R240H emphasizes the significance of alterations in the loop domain (Fig. 3), contrary to initial expectations of significant structural changes induced by the R240H mutation (Fig. 3A). Although these alterations do not severely disrupt the core beta-sheet and alpha-helix structures essential for internal dimerization or tetramerization, they may modify the protein’s dynamic behavior, especially in terms of how PERK interacts with substrates and other proteins. These findings suggest that there are other structural factors or possibly a different interaction with molecular chaperones or misfolded proteins at play with the Haplotype B and R240H variants. Such interactions might change the fine-tuned protein-protein interactions involved in ER stress detection, thereby accounting for the differences in downstream PERK signaling between variants (Fig. 4).

In summary, our research elucidates additional structural and functional consequences of PERK mutations linked to neurodegeneration. The insights support that modulation of PERK/ISR signaling is an attractive therapeutic therapy for these diseases.

## METHODS

### Comprehensive analysis of PERK variants from genomic databases

An in-depth analysis was conducted on *EIF2AK3/PERK* variants associated with WRS, tauopathy, and pLoF using three esteemed genomic databases: the Genome Aggregation Database (gnomAD version 2.1.1; Genome build: GRCh37/hg19), ClinVar (NCBI), and the European Bioinformatics Institute (EMBL-EBI) Database (IDENSG00000172071.7), up to 30 October 2023 with literature. Both the databases and relevant literature highlighted 12 WRS, 18 Tauopathy, 15 pLoF missense mutations, and 24 WRS, 13 pLoF nonsense mutations. Loss-of-function (LoF) variants are carefully classified through a detailed review of predicted pLoF variants (initially a total of 43 unique variants downloaded) that have passed additional quality control measures in the gnomAD database. In our dataset compilation, we excluded silent mutations (8 variants) and those flagged as low complexity (lc) LoF (7 variants). This analysis seeks to evaluate the potential of these variants (final 28 variants) to cause a loss of function.

### Ethnicity-Based Frequency and Population Distribution Analysis

The ethnic allele frequency for PERK SNP variants was determined by dividing the allele count by the total allele number within a population, as sourced from the Genome Aggregation (gnomAD) Database. We categorized tauopathy-associated mutations as common (≥1% frequency) and rare (<1% frequency), based on their population allele frequencies documented in gnomAD.

### Pathogenicity analysis of PERK variation

The pathogenicity of PERK clinical variants was evaluated using publicly available web server-based bioinformatics tools including PolyPhen-2, PROVEAN, MutationTaster, SIFT, and CADD (all accessed in October 2023). As a reference of the PERK gene, the NCBI Reference Sequence (gene ID 9451; Q9NZJ5.3|E2AK3_HUMAN Eukaryotic translation initiation factor 2-alpha kinase 3 OS=Homo) was used for the following analyses. PolyPhen-2 (Polymorphism Phenotyping v2) interprets amino acid substitutions by integrating sequence alignments, phylogenetics, and structural data. It provides a score ranging from 0.0 (benign) to 1.0 (probably damaging), helping to categorize the variant as ‘benign’, ‘possibly damaging’, or ‘probably damaging’. PROVEAN (Protein Variation Effect Analyzer) score is also a predictive bioinformatics tool that evaluates the potential functional consequences of amino acid substitutions within proteins. Utilizing the alignment of homologous protein sequences, it generates a score to determine the likelihood of a particular substitution being neutral or deleterious to protein function. The cutoff for PROVEAN scores was set to −2.5 for high balanced accuracy. MutationTaster predicts the functional effects of genetic variants, particularly in non-coding regions of the genome. SIFT (Sorting Intolerant From Tolerant) forecasts the potential effect of an amino acid alteration on protein functionality. It compares amino acid alignments from cognate sequences to derive a ‘SIFT score’. Scores from 0 to 0.05 are deemed ‘damaging’, while those from 0.05 to 1 are labeled ‘tolerated’. Predictions of pathological mutations lean on sequence data and neural networking. CADD (Combined Annotation Dependent Depletion) score quantifies the potential harmfulness of genetic variants by integrating diverse data types, including conservation status, functional genomics data, and population genetics insights. Typically, a scaled CADD score of 20 indicates that a variant ranks within the most deleterious top 1% across the human genome. This predictive methodology synthesizes genetic variant data with protein structural and functional characteristics, generating a lollipop graph that aggregates and averages the pathogenicity scores derived from the five bioinformatics tools, in accordance with the defined criteria presented in the following table.

### Structural modeling analysis of the PERK ER luminal domain

The reconstructed model structure of the PERK protein was visualized using the SWISS-Modeling system (https://swissmodel.expasy.org) and cross-validated with the PyMOL molecular graphics system (http://www.pymol.org). For structural modeling, we utilized the crystal structure of the human PERK luminal domain, denoted by PDB ID: 4YZY. Superimposing this reference structure with our mutation models in PyMOL, we highlighted all mismatched regions, which were depicted with circle lines for clarity.

| Value criteria of bioinformatic analysis |  |  |  |  |  | Score |
| --- | --- | --- | --- | --- | --- | --- |
|  | PolyPhen-2 | PROVEAN | Mutation Taster | SIFT | CADD |  |
| Cutoff value | $0 < \text{value} < 1$ | Cutoff = -2.5 | n/a | Cutoff = 0.05 | Cutoff = 20 | |
| Probably damaging or deleterious | $0.908 \leq$ | $< -4.1$ | disease causing | $< 0.05$ | $\geq 20$ | 4 |
| Possibly damaging | $0.446 \leq \text{value} < 0.908$ | $-4.1 \leq \text{value} < -2.5$ | | n/a | $10 \leq \text{value} < 20$ | 2 |
| Benign | $< 0.446$ | $\geq -2.5$ | polymorphism | $\geq 0.05$ | $< 10$ | 0 |

### Antibodies and chemicals

Antibodies including HSP90 (Abcam, #ab13492), PERK (Cell Signaling Technology, #3192), total (T)-eIF2α (Cell Signaling Technology, # 5324), phosphorylated (p)-eIF2α (Cell Signaling Technology, # 3398), were pre-tested to detect the expression of the targeted proteins. ER stress-inducing chemical, thapsigargin (Millipore Sigma, #T9033), was dissolved in DMSO and added to the cell culture media with a serial concentration from 1.6 pM to 1 μM. The working solution of thapsigargin was freshly prepared using stock solutions stored at −80°C.

### In vitro immunoblot analysis for ER stress sensitivity

#### Cell culture

The *PERK^-/-^*MEF cells were maintained at 37°C and 5% CO2 in complete media, composed of Dulbecco’s modified Eagle medium (Gibco™, #12430112) with 10% FBS and 1% penicillin/streptomycin (Gibco™, #15140122).

#### PERK protein expression

The *PERK^-/-^* MEF cell underwent transient transduction with four different plasmids of PERK Haplotype A and B. These contained the Dutch Alzheimer’s disease risk variant R240H and were introduced using the TransIT LT-1 Transfection reagent (Mirus Bio LLC, #MIR2020). The transfection procedure adhered strictly to the manufacturer’s guidelines. To elaborate, the *PERK^-/-^* MEF cell was plated at a density of 0.3X10^6^ cells per well in a 6-well plate. At 24 hours later, upon reaching around 70% confluency, the cells were treated with the transduction reagent. The transduction complexes were comprised of 250 µL Opti-MEM (Gibco™, #3198507), 7.5µL *LT-1* reagent (Mirus Bio LLC, #MIR2020), and 2.5 µL pDNA (having a concentration of 1µg/µL). These mixtures were left to sit at room temperature for 20 minutes before being applied to the cells, which were subsequently incubated with these complexes for an additional 24 hours.

#### Immunoblot analysis

After the aforementioned process, the transfected cells were lysed using the RIPA buffer. The protein concentrations present in the cell lysates were gauged using the BCA protein assay (Pierce). Equalized protein amounts were then loaded onto 4–15% Mini-PROTEAN TGX pre-cast gels (Bio-Rad) to be immunoblotted. The antibodies and dilutions employed included anti-HSP90 at 1:2000, PERK at 1:1000, eIF2α at 1:1000, and p-eIF2α also at 1:1000. Following an overnight incubation with the primary antibody, the membranes were rinsed in TBS containing 0.1% Tween-20. This was succeeded by an incubation period with a horseradish peroxidase-coupled secondary antibody (Cell Signaling, #7074, #7076). The immunoreactivity was then identified using the SuperSignal West chemiluminescent substrate (Pierce, #34577) and visualized via the BIO-RAD Universal Gel Molecular Imager. All western blot assays were conducted in triplicate, ensuring consistent reproducibility of the results across repeated experiments.

### Statistical analysis

For immunoblot analysis, we used Student’s t-test followed by using Prism software (GraphPad Prism, San Diego, CA). The results were used with averages of all experiments ± standard error of mean (SEM). A probability of less than 0.05 was considered statistically significant.

## Data Availability

All data produced in the present work are contained in the manuscript

## ACKNOWLEDGEMENTS

We thank Randal Kaufman for providing PERK *MEF ^-/-^* cell line.

## CONFLICT OF INTEREST

The authors declare that they have no conflicts of interest with the contents of this article.

## AUTHOR CONTRIBUTION

G.P. and J.L. conceptualized and designed the project; G.P., A.G., M.L., and K.S. collected and analyzed bioinformatics database; G.P. and K.K. performed biochemistry analysis. G.P., A.G., M.L., and J.L. co-wrote the manuscript. All authors read and approved the final manuscript.

## FUNDING AND ADDITIONAL INFORMATION

This work was supported by NIH R01NS088485 (J.L.), VA Merit I01RX002340 (J.L.), American Federation for Aging Research (J.L.), and CurePSP foundation (Kyle Kim). The content is solely the responsibility of the authors and does not necessarily represent the official views of the National Institutes of Health.

## Notes

### Competing Interest Statement

The authors have declared no competing interest.

### Funding Statement

This study was funded by NIH R01NS088485 (J.L.), VA Merit I01RX002340 (J.L.), American Federation for Aging Research (J.L.), and CurePSP foundation (Kyle Kim)

### Author Declarations

Genome Aggregation Database (gnomAD version 2.1.1; Genome build: GRCh37/hg19), ClinVar (NCBI), and the European Bioinformatics Institute (EMBL-EBI) Database (IDENSG00000172071.7)

